# Higher perivascular space volume in very premature born adults

**DOI:** 10.64898/2026.05.23.26353943

**Authors:** Niklas M. Hürter, Vijay S. Schmenger, Taylor Barda, Melissa Thalhammer, Benita Schmitz-Koep, Aurore Menegaux, Marcel Daamen, Henning Boecker, Josef Priller, Andreas Decker, Katerina Deike, Claus Zimmer, Peter Bartmann, Dieter Wolke, Benedikt Zott, Christian Sorg, Dennis M. Hedderich

**Affiliations:** Department of Diagnostic and Interventional Neuroradiology, School of Medicine and Health, TUM Klinikum Rechts der Isar, Technical University of Munich; Technical University of Munich, School of Medicine and Health, TUM-NIC Neuroimaging Center, TUM University Hospital; TUM Graduate School, TUM University, Munich, Germany; Clinical Functional Imaging Group, Department of Nuclear Medicine, University Hospital Bonn, Bonn, Germany; Department of Diagnostic and Interventional Radiology, University Hospital Bonn, Bonn, Germany; Department of Neonatology and Pediatric Intensive Care, University Hospital Bonn, Bonn, Germany; Department of Psychiatry and Psychotherapy, Klinikum rechts der Isar, School of Medicine, Technical University of Munich, Munich, Germany; Neuropsychiatry, Charité - Universitätsmedizin Berlin and German Center for Neurodegenerative Diseases (DZNE), Berlin, Germany; University of Edinburgh and UK Dementia Research Institute (UK DRI), Edinburgh, UK; German Center for Neurodegenerative Diseases, Bonn, Germany; Department of Neuroradiology, University Hospital Bonn, Bonn, Germany; Athinoula A. Martinos Center for Biomedical Imaging, Harvard Medical School, Boston, USA; Department of Psychology, University of Warwick, Coventry, UK; Warwick Medical School, University of Warwick, Coventry, UK; Institute of Neuroscience, Technical University of Munich, 80802 Munich, Germany; Munich Cluster for Systems Neurology, Technical University of Munich, 80802 Munich, Germany

**Author notes:** Corresponding author: Niklas Maximilian Hürter, Department of Diagnostic and Interventional Neuroradiology, School of Medicine and Health, TUM Klinikum Rechts der Isar, Technical University of Munich.

**Keywords:** Perivascular spaces, very preterm birth, magnetic resonance imaging, nnU-Net PVS segmentation, brain clearance

## Abstract

**BACKGROUND:** Perivascular spaces (PVS), visible on brain MRI, contribute to the brain clearance system and are associated with age and neurodegenerative disorders. While lower volumes of PVS in the forebrain’s white matter and basal ganglia have been also demonstrated in preterm-born neonates, the long-term trajectory of PVS after premature birth remains unclear. This study tests for altered PVS volumes in very preterm/very low birthweight-born (VP/VLBW) adults compared to full-term controls and explores potential associations with cognitive performance.

**METHODS:** PVS were assessed on T2-weighted MRI from 97 VP/VLBW and 89 full-term (FT) subjects at 26 years from the prospective, population-based Bavarian Longitudinal Study. PVS volume and count was based on automated nnU-Net-based segmentation. Regional PVS volumes were normalized by corresponding regional parenchyma volumes. Cognitive performance was assessed by the Wechsler Adult Intelligence Scale. MANCOVA was used for PVS group comparisons, Spearman rank correlations for testing PVS relationships with birth variables and cognitive scores.

**RESULTS:** VP/VLBW-born adults showed significantly higher normalized PVS volumes in bilateral basal ganglia (p < 0.001, partial η² = 0.096) and insula-related white matter (p = 0.001, partial η² = 0.057). In the basal ganglia, higher PVS volumes were negatively correlated with gestational age (ρ = -0.223, p = 0.030) and positively correlated with the Intensity of Neonatal Treatment Index (ρ = 0.222, p = 0.030) in the VP/VLBW group. PVS volume was not associated with IQ scores.

**CONCLUSION:** We demonstrate region-specific alterations of perivascular spaces in VP/VLBW-born adults. Data suggest that prematurity has lasting impact on the PVS.

## Introduction

Preterm birth (i.e., gestational age (GA) < 37 weeks of gestation) is associated with an increased risk for aberrant brain and neurocognitive development (Twilhaar et al., 2018, Volpe, 2009, Boardman and Counsell, 2020, Inder et al., 2023). For example, the Intelligence Quotient (IQ) after very premature birth (i.e., very preterm birth (VP i.e., GA < 32 weeks) and/or very low birth weight (VLBW < 1500g) is about 11 points lower than that of full-term (FT) peers in childhood and adulthood (Eves et al., 2021, Lacalle et al., 2023, Baranowska-Rataj et al., 2023, Breeman et al., 2015). These cognitive differences are accompanied by widespread alterations across the central nervous system, including changes in the brain’s gray matter, white matter, and cerebrospinal fluid (CSF) spaces such as enlarged ventricles (Schmitz-Koep et al., 2023, Menegaux et al., 2021, Ball et al., 2015, Rimol et al., 2019, Menegaux et al., 2020, Dimitrova et al., 2021, Skranes et al., 2007, Eikenes et al., 2011).

In addition to these structural brain alterations, the glymphatic system - the brain’s waste clearance system - has gained increasing attention in prematurity. This system represents a brain-wide network for metabolic waste removal, primarily operating through CSF circulation. This system utilizes perivascular spaces (PVS), channels surrounding penetrating brain blood vessels that communicate with the subarachnoid space, as primary conduits. Studies in mice have demonstrated that CSF enters the brain parenchyma along periarterial PVS, driven by pulsatile movements of the arterial vessel wall (Holstein-Rønsbo et al., 2023, Rasmussen et al., 2022, Iliff et al., 2012, Potter et al., 2015, Wardlaw et al., 2020, Kwee and Kwee, 2007, Jessen et al., 2015, Abbott et al., 2018, Plog and Nedergaard, 2018, Mestre et al., 2018). Importantly, PVS characteristics visible on magnetic resonance imaging (MRI) may serve as markers of glymphatic system function, with enlarged PVS potentially indicating disturbed CSF transport and clearance (Wardlaw et al., 2020). In humans, enlarged PVS can be detected by T1- or T2-weighted MRI together with automated PVS segmentation (Wardlaw et al., 2020, Kwee and Kwee, 2007). Such PVS alterations have been documented in normal aging and various neurological conditions including Alzheimer’s disease, cerebral small vessel disease, and Parkinson’s disease (Potter et al., 2015, Wang et al., 2022, Menze et al., 2024). A small number of neonatal studies have begun to investigate PVS alterations in prematurity, reporting inconsistent findings across brain regions (Kim et al., 2023b, Lin et al., 2024, Meinhold et al., 2025). Whether such alterations persist into adulthood following preterm birth remains unknown.

We hypothesized aberrant PVS volumes in adults born VP/VLBW, which link with perinatal factors such as lower GA or BW or the degree of neonatal neurological complications. Furthermore, we explored whether potential PVS alterations were related with aberrant neurocognitive performance as measured by IQ. To address these hypotheses, we used T2-weighted MRI and machine learning-based automated segmentation to investigate differences in PVS volumes between VP/VLBW-born and term-born adults at 26 years of age. Group comparisons and relationships with perinatal variables or IQ measures were performed by MANCOVA and correlation analysis.

## Methods

### Participants

Our participants were part of the Bavarian Longitudinal Study (BLS), a geographically defined whole population-based study of VP/VLBW-born and FT-born infants born in southern Bavaria, Germany, between Jan 1, 1985 and March 31, 1986 (previously described in, for example, (Schmitz-Koep et al., 2023, Thalhammer et al., 2024a, Schmitz-Koep et al., 2021, Schmitz-Koep et al., 2020, Thalhammer et al., 2024b, Hedderich et al., 2020, Breeman et al., 2015). Initially 682 VP/VLBW infants (defined as GA < 32 weeks and/or BW < 1500g) and 916 healthy term infants (GA > 37 weeks) were included. At age 6, the control group was strategically reduced to 350 children through random selection, maintaining comparability with the VP/VLBW group regarding sex and family socioeconomic status. By age 26, 411 VP/VLBW and 308 FT participants were eligible for follow-up assessment in general, from whom 212 participants (101 VP/VLBW and 111 FT) underwent MRI at either the Department of Radiology, University Hospital of Bonn (n = 67) or the Department of Neuroradiology, Klinikum rechts der Isar, Technical University of Munich (n = 145). The study was carried out in accordance with the Declaration of Helsinki and was approved by the local ethics committee of the Klinikum rechts der Isar, Technical University of Munich, and the University Hospital Bonn. All study participants gave written informed consent.

They received travel expenses and payment for participation.

After excluding 25 cases with incomplete or impaired MRI data and one FT subject after controlling for outliers (for more detail see below), the final study sample consisted of 186 participants (97 VP/VLBW and 89 FT) (Table 1).

**Table 1:** Clinical characteristics of VP/VLBW and full- term-born adults.

| **Variable** | **VP/VLBW (n=97)** | **FT (n=89)** | **p-value** |
| --- | --- | --- | --- |
| **Sex, n (%)** |  |  | 0.704 <sup>3</sup> |
| - Male | 54 (55.7%) | 52 (58.4%) |  |
| - Female | 43 (44.3%) | 37 (41.6%) |  |
| **Age, years** | 26.74 ± 0.60 | 26.97 ± 0.75 | 0.027 <sup>2</sup> |
| **Gestational age, weeks** | 30.48 ± 2.09 | 39.70 ± 1.09 | <0.001 <sup>2</sup> |
| **Birth weight, g** | 1326.03 ± 316.20 | 3374.93 ± 438.06 | <0.001 <sup>1</sup> |
| **Hospital stay, days** | 72.28 ± 26.54 | 6.73 ± 2.39 | <0.001 <sup>2</sup> |
| **Intensity of Neonatal Treatment Index** | 11.63 ± 3.78 | NA | - |
| Full-Scale IQ | 94.60 ± 12.63 | 102.91 ± 11.79 | <0.001 <sup>1</sup> |

### Birth-related variables

GA was estimated by maternal reports and ultrasound diagnostics during pregnancy. In cases in which the two results differed for more than two weeks, it was estimated using the Dubowitz method (Dubowitz et al., 1970). BW, duration of hospitalization (DH), and Intensity of Neonatal Treatment Index (INTI), quantifying duration and intensity of medical treatment after birth, were obtained from the neonatal records (Gutbrod et al., 2000). Daily assessments of care level, respiratory support, feeding dependency, and neurological status (mobility, muscle tone, and neurological excitability) were performed. Each of the 6 variables was scored on a 4-point rating scale (0–3) by the method of Casaer and Eggermont (Casaer P, 1985). The INTI was computed as the mean score of daily ratings during the first 10 days of life or until a stable clinical state was reached (total daily scores < 3 for 3 consecutive days), depending on which occurred first, ranging from 0 (best state) to 18 (worst state).

### Cognitive performance assessment

Global cognitive performance was assessed using a short version of the Wechsler Adult Intelligence Scale (WAIS-III; German adaptation: “Wechsler Intelligenztest für Erwachsene”, WIE) (von Aster M, 2006). Testing was conducted prior to MRI examination by trained psychologists who were blinded to participant group status (Breeman et al., 2015, Eryigit Madzwamuse et al., 2015). Full-scale IQ estimates were derived from these assessments.

### MRI data acquisition, preprocessing, and definition of regions-of-interest

MRI data acquisition (previously described in (Schmitz-Koep et al., 2021, Hedderich et al., 2020) was performed at both sites (Bonn and Munich) using Philips Achieva 3T TX or Philips Ingenia 3T systems, each equipped with an 8-channel SENSE head coil. Subject distribution across scanners was as follows: Bonn Achieva 3T (5 VP/VLBW, 12 FT), Bonn Ingenia 3T (33 VP/VLBW, 17 FT), Munich Achieva 3T (60 VP/VLBW, 65 FT), and Munich Ingenia 3T (3 VP/VLBW, 17 FT). Sequence parameters (see below) were kept identical across all scanners. Quality assurance included regular phantom scans by MRI physicists at both sites to ensure within- scanner signal stability. Signal-to-noise ratio showed no significant differences between scanners (one-way ANOVA with factor ’scanner-ID’; F (3.182) = 1.84, p = 0.11). Scanner-specific dummy variables were included as covariates-of-no-interest to account for potential scanner effects. A high-resolution T1-weighted 3D-MPRAGE sequence (TI=1300ms, TR=7.7ms, TE=3.9ms, flip angle=15°, field of view=256 mm × 256 mm, reconstruction matrix=256×256, reconstructed isotropic voxel size=1 mm^3^) and a high-resolution T2-weighted 3D sequences (TR = 2500 ms, TE = 364 ms, flip angle = 90°, field of view = 256 × 256 mm acquisition matrix: 256 × 256, reconstruction matrix: 512 × 512, echo train length = 120, resulting in a reconstructed isotropic voxel size of 0.5 × 0.5 × 0.5 mm were acquired for each participant.

From the initial cohort of 212 subjects, 12 were excluded due to missing T2-weighted images. For the remaining 200 subjects, images were converted from DICOM to NIfTI format using the MRIcron dcm2nii conversion tool *(Li et al., 2016)*, followed by brain extraction using FSL’s bet (version 6.0, fractional intensity threshold = 0.3). The resulting skull-stripped images were registered to MNI152 standard space using NiftyReg’s reg_aladin (version 1.5.58, http://cmictig.cs.ucl.ac.uk/wiki/index.php/NiftyReg) (Modat et al., 2014). This registration employed a combined rigid and affine transformation with cubic interpolation (Example of the used command: reg_aladin -ref MNI152_T1_1mm_brain.nii.gz \ -flo <subject_T2.nii.gz> \ -aff <output_affine.txt> \ - res <registered_T2.nii.gz>), using the 1 mm³ isotropic MNI152 T1-weighted skull-stripped template as reference. During this registration process, images were resampled from native 0.5 mm³ to 1 mm³ isotropic resolution (for more details see below and Supplementary Methods, ’Comparison of MRI Input Data’).

To prepare a regional analysis of PVS, we utilized FreeSurfer’s recon-all *white matter parcellation* (wmparc.mgz, FreeSurfer version 7.3.2). Wmparc parcellates the white matter (WM) in relation to foregoing canonical grey matter parcellation based on Desikan-Killiany atlas. A complete list of wmparc codes for each region is provided in Supplementary Table S1. Individual wmparc labels were grouped into larger anatomical WM regions based on standard neuroanatomical boundaries. This coarse parcellation included frontal, parietal, occipital, temporal, insular, cingulate and unsegmented WM, as well as basal ganglia. Deep white matter structures were classified as unsegmented if assignment to specific cortical regions is not possible (see Figure 1 for spatial outline of WM regions used in this study).

**Figure 1:**
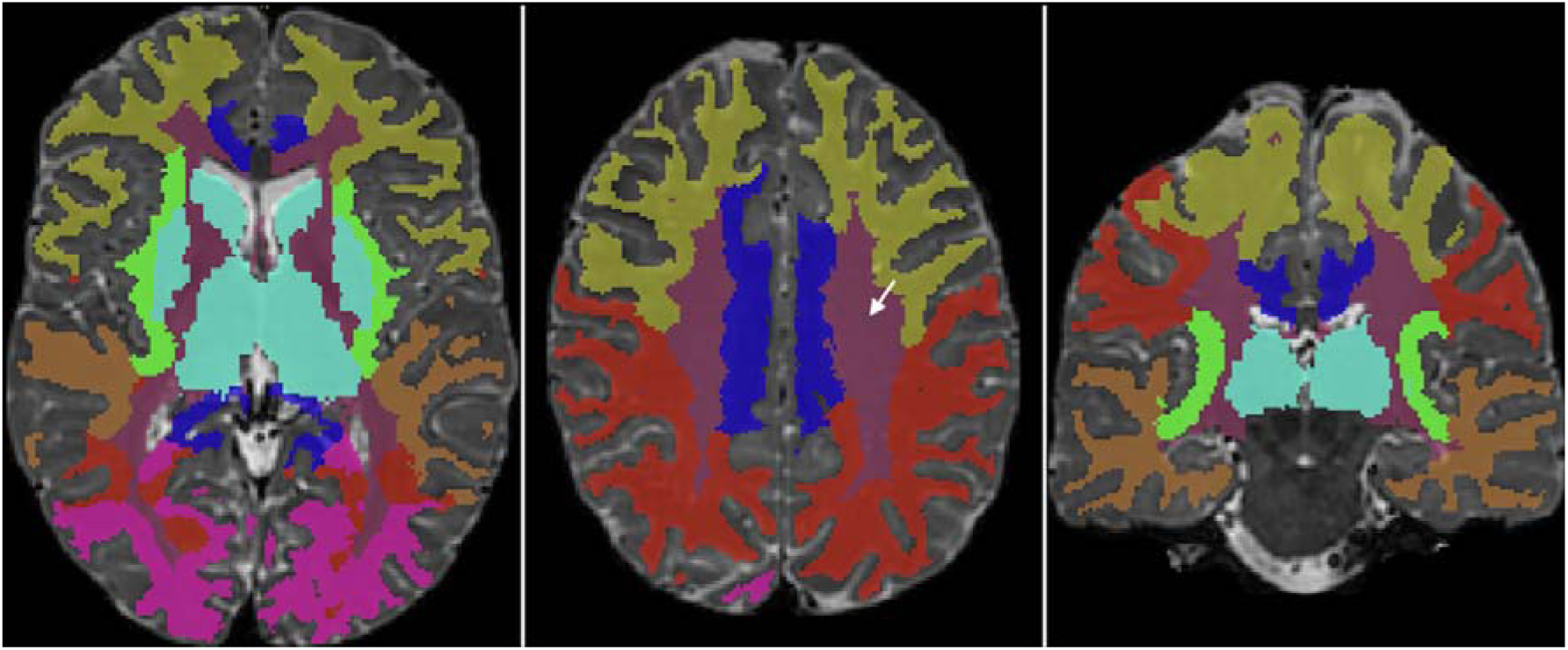
Regional segmentation of white matter and basal ganglia based on FreeSurfer’s recon-all white matter parcellation tool (wmparc.mgz, FreeSurfer version 7.3.2). Parcellated regions are grouped in frontal, parietal, occipital, temporal, insular, cingulate and unsegmented WM, as well as basal ganglia.

**Figure 2:**
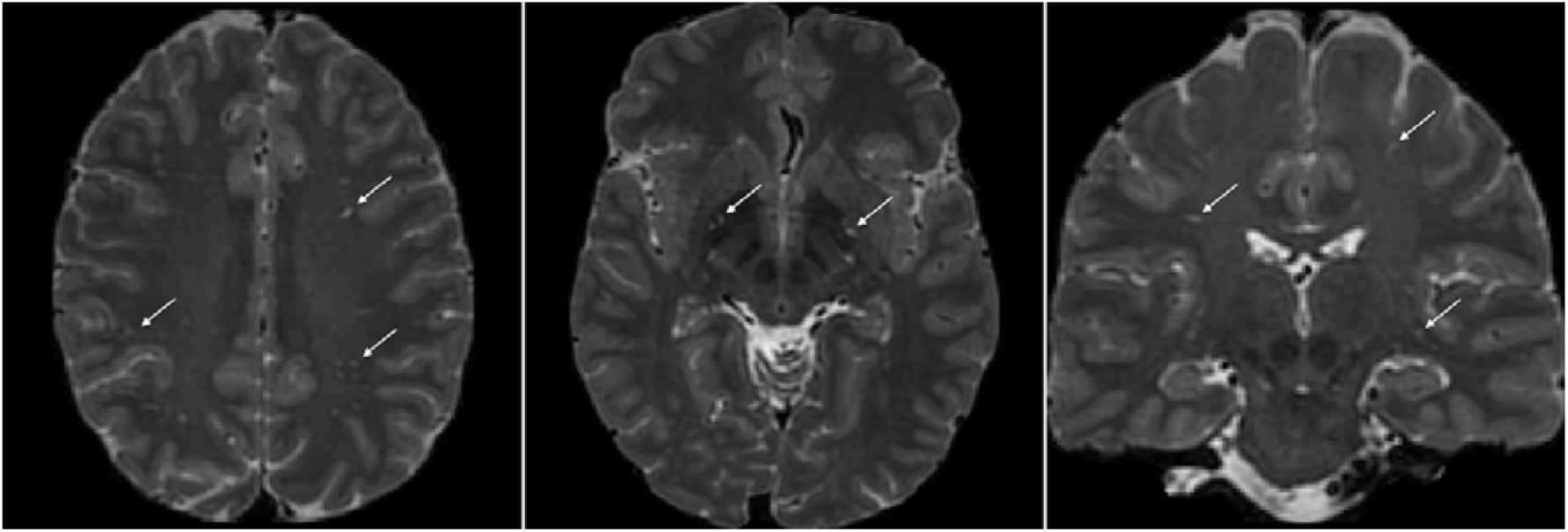
PVS in T2-weighted MRIs. Left) PVS in the centrum semiovale, axial view. Middle) PVS in the basal ganglia. Right) In the basal ganglia, coronal view. White arrows indicate representative PVS in all images.

**Figure 3:**
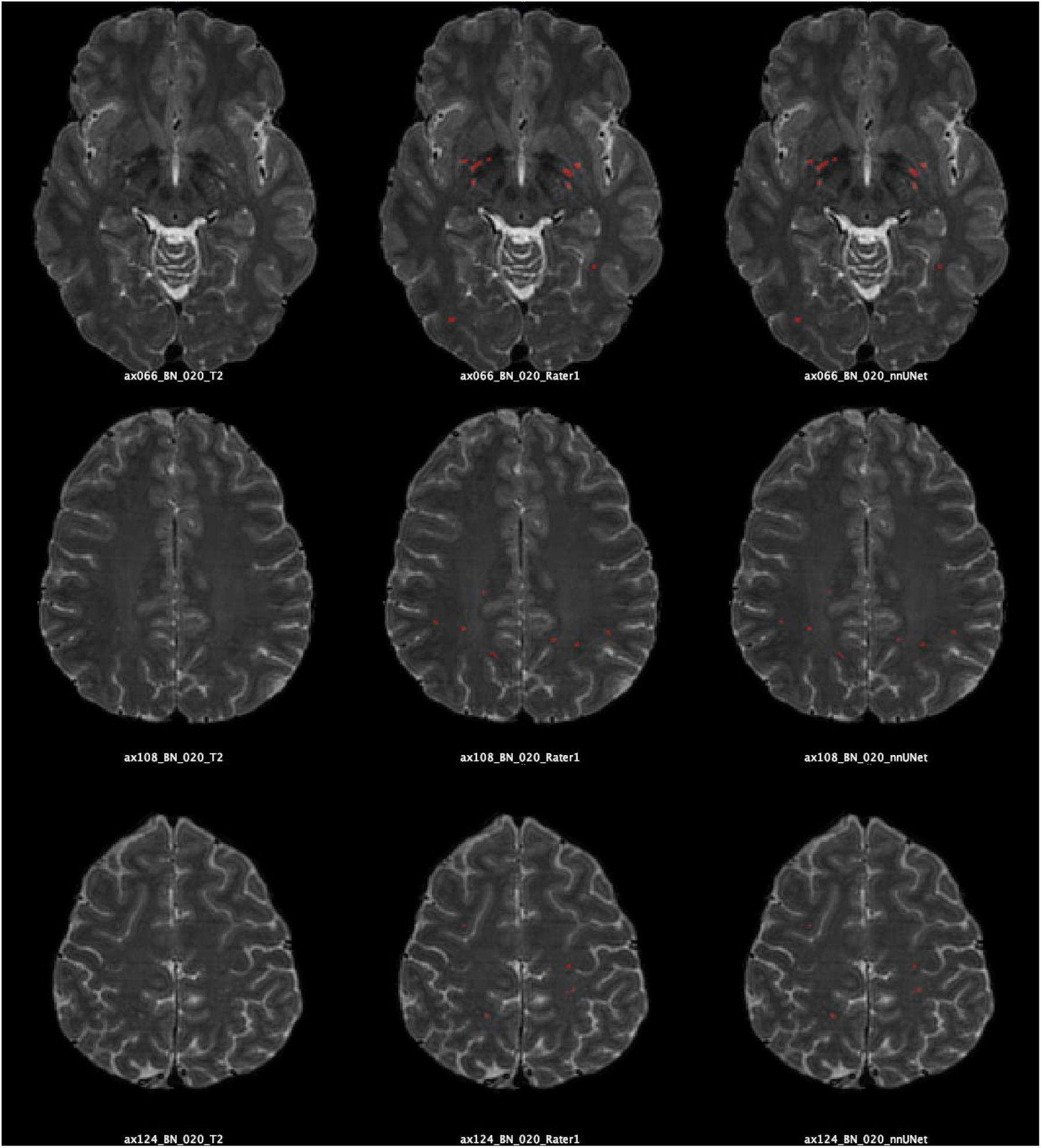
Comparison of manual and automated PVS segmentation. Representative example of automated PVS segmentation of one subject across three planes. Left column: Native T2-weighted image for three planes. Middle column: Manual segmentation by Rater 1 (red overlay). Right column: Automated nnU-Net segmentation (red overlay). Examples were selected to represent typical algorithm performance (DSC = 0.637 ± 0.037 across the cohort).

**Figure 4:**
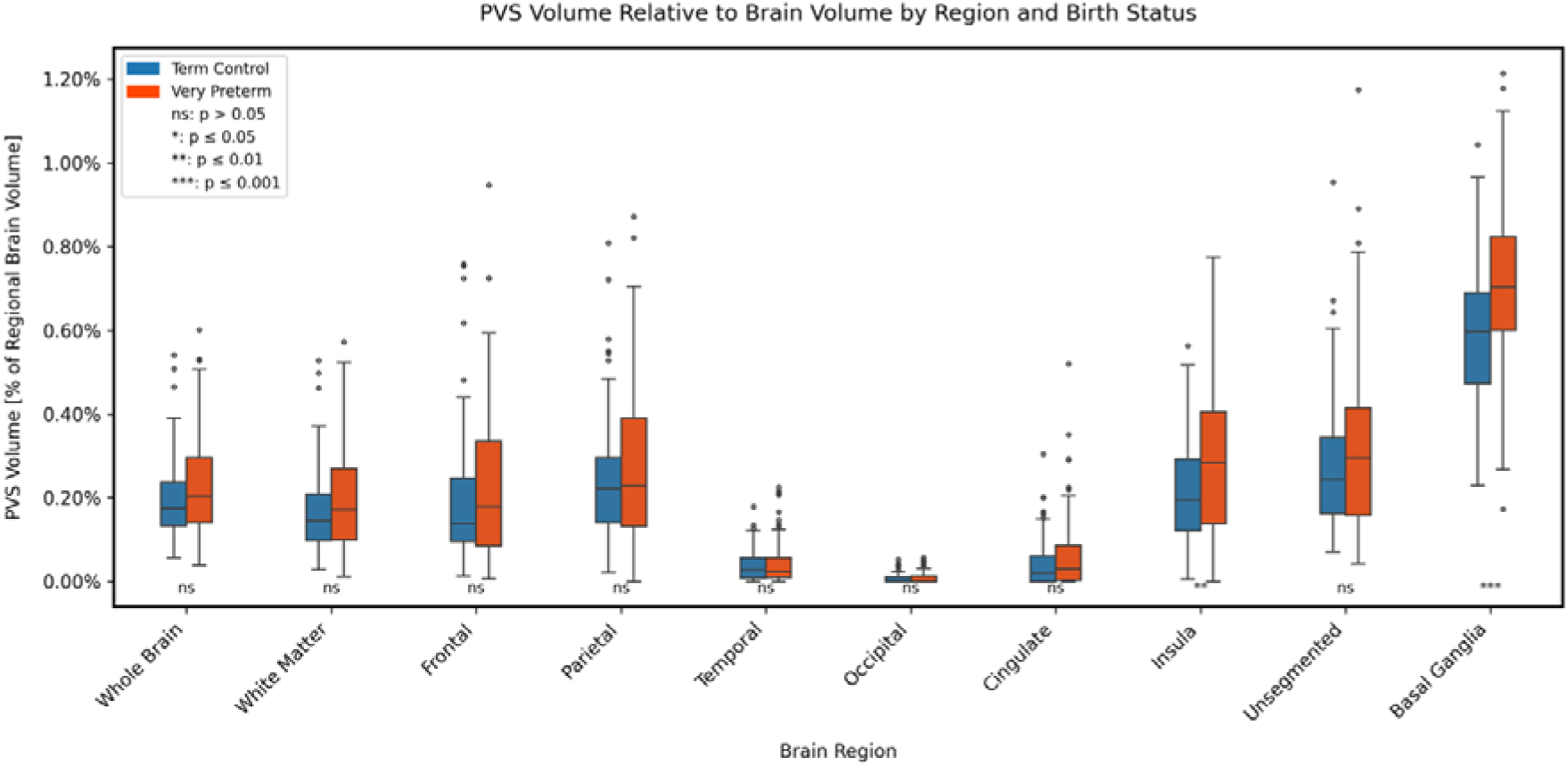
Regionally normalized PVS volumes. Regional normalized PVS volumes in VP/VLBW and full-term-born adults. Significant group differences are marked by stars. MANCOVA with LSD post-hoc test. * p < 0.05, ** p < 0.01, *** p < 0.001).

**Figure 5:**
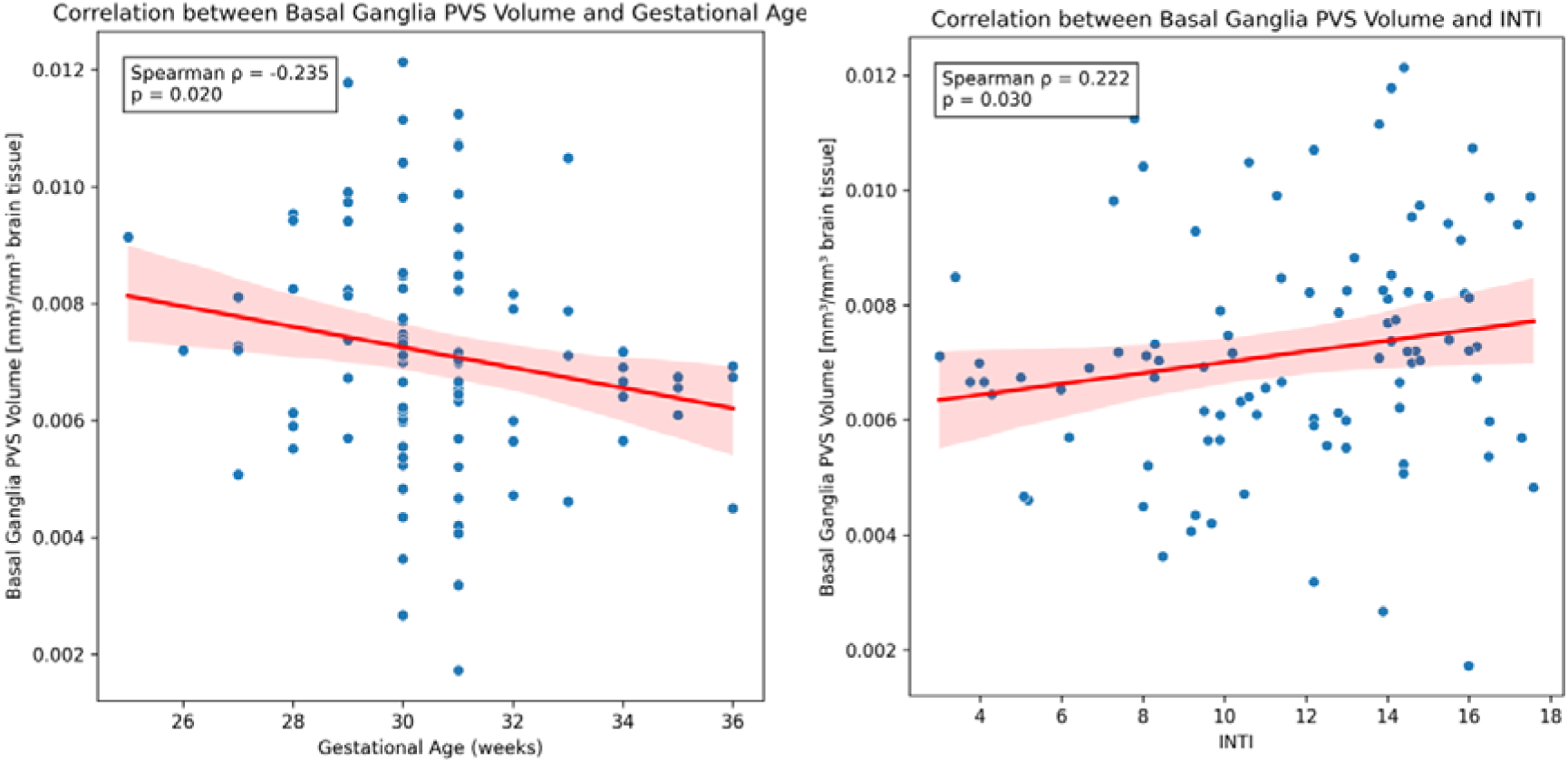
Association between regionally normalized PVS volumes and birth variables in VPT/VLBW adults. Spearmen correlation analysis. Birth variables are gestational age and the intensity of neurological treatment index INTI.

Visual quality control of these pre-processed MRIs was performed independently by two trained raters (NH, TB). Images were excluded from further analysis if they met any of the following criteria: presence of significant motion artifacts (including blurring, ghosting, or distortions) that would affect PVS quantification (6 subjects excluded), unsuccessful registration to MNI space (2 subjects excluded), or failed mask generation during FreeSurfer processing (5 subjects excluded). The final study sample comprised 186 participants (97 VP/VLBW and 89 FT-born adults) after one additional subject was excluded as an outlier for PVS volume distribution.

### PVS: segmentation approach and PVS outcomes

There are several automated segmentation approaches that differ in segmentation algorithm and MRI input modality for PVS segmentation. To identify the optimal protocol for our dataset, we systematically compared following approaches: Regarding segmentation algorithm, we compared WPSS (Weakly Supervised Perivascular Space Segmentation) and nnU-Net. WPSS is a deep learning method specifically developed for PVS segmentation, combining Frangi-filter-derived (Frangi et al., 1998) geometric priors with convolutional neural network-based feature learning (Lan et al., 2023). nnU-Net is a self-configuring deep learning framework that has demonstrated state-of-the-art performance across a broad range of medical image segmentation tasks (Isensee et al., 2021).

Regarding imaging modality, we compared T2-weighted images with Enhanced Perivascular Contrast (EPC) images, the latter generated by voxel-wise division of T1- by T2-weighted images to enhance PVS-to-tissue contrast (Sepehrband et al., 2019). Since EPC generation requires both modalities at identical resolution and our T1-weighted images were acquired at 1 mm³, both modalities were standardized to 1 mm^3^ isotropic resolution for this comparison.

Based on this comparison, we selected the combination of T2-weighted imaging and nnU-Net as our final segmentation pipeline, as it yielded the highest segmentation performance on our dataset. A full and detailed description of our methodological pre-study is reported in the Supplementary Methods.

### PVS segmentation: nnU-net-based segmentation and PVS outcomes

#### Manual segmentation and ground truth

On T2-weighted MRI, we identified PVS based on established imaging criteria including their typical appearance as fluid-filled tubular structures. Most PVS are visible in the basal ganglia around lenticulostriate arteries and in the WM centrum semiovale along medullary arteries. Based on their orientation to the imaging plane, PVS present either as linear (when parallel) or round/ovoid structures (when perpendicular), appearing hyperintense on T2w images.

To develop and validate an automated PVS segmentation pipeline applicable to our full cohort, we first established ground truth through manual PVS segmentation in a representative subsample, ensuring balanced distribution across all four scanners, which served as training data for the present machine learning approach. Manual PVS annotation was performed on T2w images in all three planes (i.e. axial, sagittal, and coronal) using ITK-Snap (version 4.0.2) by a trained medical evaluator from neuroradiology (Rater 1, NH) of 40 subjects. For validation purposes, a trained neuroscientist (Rater 2, TB) independently annotated a randomly selected subset of 20 subjects (50%).

#### nnU-net-based segmentation

For automated PVS segmentation, we used a nnU-Net approach (Isensee et al., 2021). Manual annotations from Rater 1 on 40 subjects served as ground truth for algorithm training. The nnU-Net implementation (version 2, February 2024) utilized a NVIDIA GeForce GTX 3060 (12GB VRAM) with 3D full-resolution configuration. NnU-Net auto-configured parameters included batch size=2, patch size=128x128x128, and learning rate ranging from 0.01 to Initial training used 32 subjects for training and 8 for validation, randomly selected, followed by five-fold cross-validation on all 40 subjects to obtain robust performance estimates. Segmentation performance was evaluated using Dice Score Coefficient (DSC), Sensitivity (SEN), and Positive Predictive Value (PPV), calculated based on true positives (TP), false positives (FP), and false negatives (FN) as defined in the formulas below. Reported values represent mean performance across cross-validation folds.

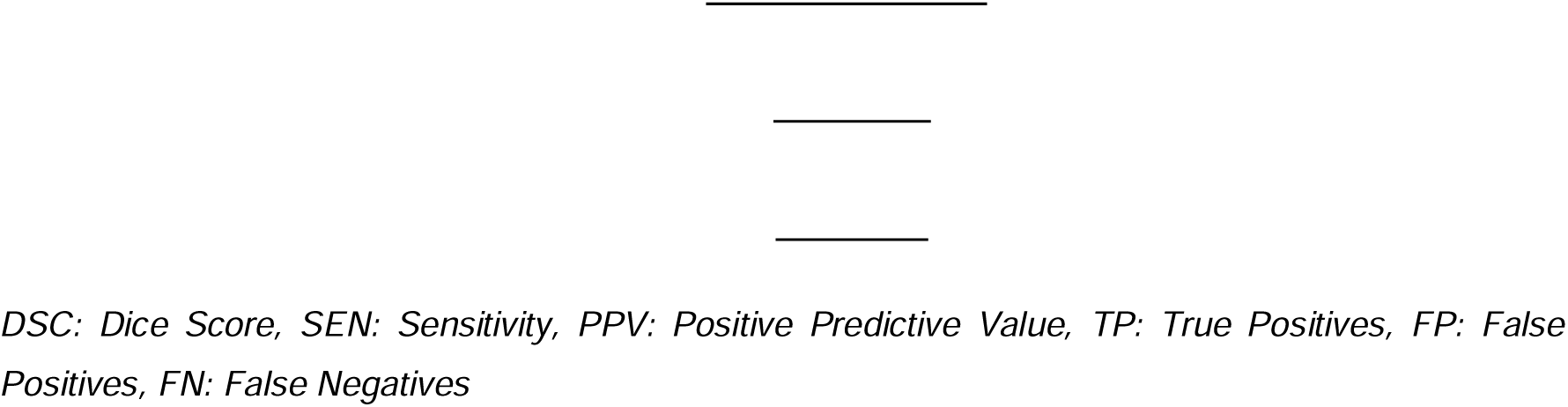

After training, the algorithm achieved a DICE score of 63.7% compared to Rater 1, close to the inter-rater DICE score of 53.8% between Rater 1 and Rater 2, indicating that the automated segmentation performed within the range of human rater variability and corresponds to human-level segmentation performance given the available image quality. Performance metrics represent mean values across three-dimensional volumetric evaluation of all cross-validation folds. Regional analysis showed stronger performance in the basal ganglia (DSC = 0.716, sensitivity = 0.665, PPV = 0.791) compared to white matter (DSC = 0.611, sensitivity = 0.525, PPV = 0.764). This performance is comparable to other recent nnU-Net implementations for PVS segmentation on 3T MRI data. Cai et al. (Cai et al., 2024) reported nnU-Net DSC of 68% on T2-weighted 3T data, while the VALDO challenge (Sudre et al., 2024) reported 61.9% DSC for nnU-Net-based PVS segmentation.

To promote reproducibility and facilitate application to other cohorts, we have made the trained model weights publicly available (https://github.com/vijaysebastianS/nnU-Net-for-PVS-Segmentation-on-3T-T2w-MRI).

#### PVS outcomes

PVS burden was quantified using two metrics: volume and count. PVS volume was calculated as total volume of annotated PVS in cubic millimeters (mm³) per brain region. PVS count was determined through three-dimensional cluster analysis, where interconnected voxels (sharing either faces, edges, or vertices) were defined as single PVS entities. The final count represented the number of distinct PVS clusters per brain region. While count provides valuable information about PVS distribution, it may underestimate burden in cases where adjacent PVS have merged or where individual spaces occupy a larger volume, making PVS volume the preferred metric for capturing prematurity-induced alterations.

PVS volumes are known to strongly correlate with brain size in healthy individuals (Barisano et al., 2021), making normalization necessary to enable valid inter-subject comparisons. We calculated regional PVS volume fraction by dividing the total PVS volume within each anatomical region by the corresponding regional tissue volume. This normalization approach, established by Zong et al. (Zong et al., 2016) and adopted in subsequent quantitative PVS studies (Sepehrband et al., 2019, Sepehrband et al., 2021, Barisano et al., 2022), accounts for both global brain size differences and regional heterogeneity in PVS distribution. To verify robustness, we performed control analyses using alternative normalization strategies (see Supplementary Table S9).

### Statistical analysis

All statistical analyses were performed using IBM SPSS Version 29 (IBM Corp). Statistical significance was assessed at p < 0.05, and correction of multiple testing was based on false discovery rate (FDR) procedures. Extreme outliers, defined as values exceeding 3 standard deviations from the mean in the three main regions of interest (whole brain, white matter, and basal ganglia) were excluded. This outlier analysis led to the exclusion of one FT subject.

#### Group comparisons

Group comparisons of normalized PVS metrics were conducted using a Multivariate Analysis of Covariance (MANCOVA), with PVS values of different regions as dependent variables and group status as the independent variable, while controlling for sex, age and scanner type as covariates of no interest. Analyses were performed both on a whole-brain level and for specific regions. For regions showing significant group differences, additional hemisphere-specific analyses were conducted to evaluate potential laterality of effects. To control the findings for potential methodological decisions, we repeated analyses for different normalization approaches: (1) normalization via division by estimated total intracranial volume (eTIV) instead of regional volume, (2) statistical normalization by including eTIV volume as covariate. The results were corrected for multiple comparisons using the Benjamini–Hochberg procedure (Benjamini Y, 1995). Statistical significance was defined as p<0.05, FDR-corrected.

#### Correlation analysis - birth variables

To examine whether PVS volume differences were specifically related to preterm birth, we additionally performed Spearman correlation analyses for the significant regions with birth-related variables (i.e., GA, BW, DH, INTI) within the VP/VLBW group.

#### Correlation analysis – cognitive performance

Finally, to explore whether PVS volume differences were related to cognitive performance, we performed Spearman correlation analyses with full-scale IQ.

PVS count analyses followed a similar analytical framework but served as secondary analyses to validate the primary volume-based findings.

## Results

### Demographic and clinical characteristics

The demographic and clinical characteristics of the study groups are presented in Table 1. The VP and FT groups showed comparable distribution regarding sex (44.3% vs 41.6% female, p = 0.704), and small but significant difference in age at scan time (VP = 26.74 ± 0.60 vs FT = 26.97 ± 0.75 years, p = 0.027). As expected by the study design, VP participants had significantly lower GA (30.48 ± 2.09 vs 39.70 ± 1.09 weeks, p < 0.001) and BW (1326.03 ± 316.20 vs 3374.93 ± 438.06 g, p < 0.001) compared to term-born participants. Cognitive assessment revealed significantly lower performance in the VP group as measured by full-scale IQ (VP = 94.60 ± 12.63 vs FT = 102.91 ± 11.79, p < 0.001).

### Regional PVS volume differences between VP/VLBW and FT Adults

Multivariate analysis of variance revealed significant overall group differences in PVS volumes (Pillai’s Trace = 0.211, F(10.172) = 4.612, p < 0.001). VP/VLBW adults showed higher regionally normalized PVS volumes in the basal ganglia (p < 0.001, partial η² = 0.096), the insula-related WM (p = 0.001, partial η^2^ = 0.057), and cingulate WM (p = 0.039, partial η² = 0.023) (see Table 2). After FDR correction for multiple testing, both the basal ganglia and insula differences remained significant.

**Table 2:** Group differences for regional PVS volumes. * indicates p<0.05; ** indicates p_FDR_<0.05.

| Region | VP/VLBW Mean (SD) | Full-Term Mean (SD) | p-value | FDR-correction cut off |
| --- | --- | --- | --- | --- |
| whole brain | 0.002305 (0.001217) | 0.001988 (0.000953) | 0.056 | 0.02 |
| whole WM | 0.001951 (0.001285) | 0.001680 (0.001000) | 0.121 | 0.03 |
| frontal WM | 0.002298 (0.001853) | 0.001908 (0.001549) | 0.132 | 0.035 |
| parietal WM | 0.002788 (0.001970) | 0.002487 (0.001458) | 0.250 | 0.045 |
| temporal WM | 0.000415 (0.000518) | 0.000379 (0.000378) | 0.599 | 0.05 |
| occipital WM | 0.0000953 (0.000157) | 0.0000680 (0.000112) | 0.180 | 0.04 |
| cingulate WM | 0.000645 (0.000855) | 0.000418 (0.000572) | 0.039* | 0.015 |
| insula WM | 0.002943 (0.001828) | 0.002160 (0.001323) | 0.001** | 0.01 |
| unsegmented WM | 0.003233 (0.001961) | 0.002787 (0.001588) | 0.094 | 0.025 |
| basal ganglia | 0.007129 (0.001999) | 0.006000 (0.001575) | <0.001** | 0.005 |

To test the reliability and plausibility of this main finding, we conducted the following control analyses. First, PVS volumes were normalized by division through eTIV (see Table S9); higher eTIV-normalized PVS volumes were observed in VP/VLBW adults for the insula-related WM (p = 0.002, partial η^2^ = 0.052) and basal ganglia (p = 0.018, partial η^2^ = 0.030), supporting our main finding. Second, rather than normalizing by division, eTIV was included as an additional covariate in the MANCOVA model (see Table S9); this statistical normalization likewise yielded higher PVS volumes in VP/VLBW adults for the insula-related WM (p < 0.001, partial η^2^ = 0.069).

Then we tested the plausibility of our PVS volume findings by comparing PVS volumes we detected with PVS volumes of previous recently published studies in healthy adults (Barisano et al., 2021, Kim et al., 2023a). For white matter, our cohort (VP/VLBW and FT) showed a mean absolute volume of 886 ± 582 mm^3^, comparable to the 758 ± 485 mm³ reported by Kim et al. in healthy young adults, while Barisano et al. reported substantially higher absolute values (5,029 ± 2,153 mm^3^). When expressed as eTIV-normalized fraction, our values likewise closely matched Kim et al. (0.055 ± 0.035% vs. 0.056 ± 0.035%), whereas Barisano et al. reported a substantially higher regional normalized fraction (1.14 ± 0.43%). For basal ganglia, our absolute volume of 233 ± 62 mm^3^ was 2.7-fold higher than the 85 ± 31 mm^3^ reported in Kim et al.’s healthy controls, with corresponding eTIV-normalized fractions of 0.015 ± 0.004% vs. 0.006 ± 0.002%. Thus, our PVS volumes were plausible in comparison to those of Kim et al..

Then, we controlled our main finding by replacing PVS volume as outcome by PVS count. For normalized PVS counts, significant group differences were observed in basal ganglia (p < 0.001, partial η^2^ = 0.060), insula -related WM (p = 0.010, partial η^2^ = 0.036), and cingulate WM (p = 0.030, partial η^2^ = 0.026), supporting our main finding of increased PVS volumes in the same regions (the complete results are provided in the supplementary Table S13) (iv) Finally, we asked whether our main finding might be biased by hemispheric different PVS volume alterations. Subsequent analysis of PVS volumes of each hemisphere, respectively, revealed that in the VP/VLBW group, significantly higher PVS volumes were found in both right and left basal ganglia (right: p < 0.001, partial η^2^ = 0.082; left: p = 0.001, partial η^2^ = 0.056) and insula -related WM regions (right: p = 0.026, partial η^2^ = 0.027; left: p = 0.001, partial η^2^ = 0.060).

Thus, these control analyses support our main finding of higher PVS volumes in very premature adults for the basal ganglia and the white matter around the insulae.

### Relationship between PVS volumes and birth variables

To further ensure that higher PVS volumes of very premature born adults is indeed related to preterm birth, we examined correlations between PVS volumes and birth-related variables within the VP/VLBW group. We observed significant correlations of basal ganglia PVS volumes with both GA (Spearman correlation: ρ = -0.235, p = 0.020) and INTI (ρ = 0.222, p = 0.030), suggesting that basal ganglia PVS volume increases in preterm born adults are due to premature birth and its complications.

### No relationship between PVS volumes and cognitive performance

Finally, we investigated potential associations between PVS volume increases in the basal ganglia and insula and impaired cognitive performance of VP/VLBW-born adults using Spearman correlation. As proxy for cognitive performance, we used Full-Scale IQ. Preterm born adults had lower IQ (94.60 ± 12.63 vs. 102.91 ± 11.79; p<0.001). We observed that IQ scores were not significantly correlated with higher PVS volumes of insula (Spearman ρ = 0.170, p = 0.102) and basal ganglia (Spearman ρ = -0.047, p = 0.653).

## Discussion

Using T2w-MRI in very premature and full-term born subjects, we observed higher PVS volumes in the basal ganglia and insula-related WM of VP/VLBW-born adults. To the best of our knowledge, this study provides the first evidence for PVS alterations in premature born adults. The observed regional pattern of PVS alterations in prematurity aligns with current knowledge about the selective vulnerability of both insula and basal ganglia after preterm birth (Boardman et al., 2006, Srinivasan et al., 2007, Nosarti et al., 2014, Nosarti et al., 2008). Thus, the present findings support the hypothesis that prematurity has sustained effects on the PVS particularly in basal ganglia and insula-related WM with potential impact on glymphatic brain clearance.

The present finding of higher PVS volumes in basal ganglia correlating with INTI and anti-correlating with GA suggests that preterm birth impacts PVS development. This is confirmed by the results of Meinhold et al. (Meinhold et al., 2025), who reported elevated PVS counts in the centrum semiovale of very preterm-born neonates. In contrast, Kim et al. (Kim et al., 2023a) found lower basal ganglia PVS volumes in preterm neonates – a discrepancy that may reflect developmental trajectories shifting between the neonatal period and adulthood, though this cannot be resolved by our cross-sectional design. Of note, higher PVS volumes are also observed in aging and neurodegeneration (Menze et al., 2024, Huang et al., 2021, Potter et al., 2015, Wang et al., 2022), where they have been associated with impaired glymphatic clearance (Wardlaw et al., 2020, Plog and Nedergaard, 2018). By analogy, the higher PVS volumes in VP/VLBW-born adults observed here may reflect altered flow and clearance capacity in the CSF-glymphatic system following preterm birth.

The discrepancy between our PVS volumes and those reported in the literature merits discussion, as it highlights important methodological considerations in quantitative PVS research. Our white matter values closely match Kim et al. (Kim et al., 2023a) (886 vs. 758 mm³; 0.055% vs. 0.056% ICV-normalized), validating our technical approach when methodologies are similar. In contrast, Barisano et al. (Barisano et al., 2021) reported approximately 5-fold higher values. It is likely reflecting differences in analytical resolution (native 0.7mm vs. our 1mm standardization) and segmentation methodology (Frangi filtering with broad scale parameters optimized for high sensitivity vs. our conservative deep learning approach trained on manual annotations with inherent variability, DSC = 53.8%). Importantly, while absolute volumetric calibration varies across studies depending on technical and analytical choices, the internal validity of our group comparisons remains robust: the critical finding is the significant difference between VP/VLBW and control groups within our cohort, measured with consistent methodology applied equally to both groups.

The lack of correlation between higher PVS volumes and cognitive performance seems plausible if PVS volumes are connected to functions of the glymphatic system. Their alterations might be more relevant for long-term brain health (Passiak et al., 2019) and consequences might occur only later in life, rather than impacting current cognitive function in young adulthood. Indeed, at age 26, potential downstream effects of impaired brain clearance might not yet be apparent in cognitive performance, though they could potentially influence age-related processes later in life.

## Limitations and strengths

Several methodological strengths support the validity of our findings: (i) Our use of advanced machine learning techniques for PVS segmentation, which have been extensively validated against other segmentation approaches, provides robust quantification of PVS volumes. (ii) Our population-based approach with a cohort size that is considerable for this type of investigation, combined with careful control analyses, further strengthen the conclusions drawn.

Our study has also some limitations. (i) The cross-sectional nature of our MRI study, due to the fact that these are the first MRI scans of this population, prevents direct assessment of PVS development over time. (ii) Additionally, while we controlled for scanner effects in our analyses, the use of multiple scanners introduces potential variability that future single-scanner studies might avoid. (iii) Furthermore, accurate PVS segmentation at 3T MRI (instead of 7T MRI) remains challenging due to the small size of these structures, limited contrast resolution, and their complex anatomical distribution along blood vessels, which might affect the precision of volumetric measurements despite using validated techniques. (iv) Another important limitation concerns potential confounding factors known to influence PVS volume in healthy adults. Quantitative studies have shown consistent associations of body mass index with white matter PVS volume and demonstrated that circadian rhythm and time of day can affect measurements through sleep–wake–dependent glymphatic variations (Barisano et al., 2021). Genetic factors also account for inter-individual variability in PVS burden (Barisano et al., 2021), while cardiovascular risk factors such as hypertension are particularly associated with enlarged PVS in the basal ganglia (Wardlaw et al., 2020, Potter et al., 2015). Age remains one of the strongest predictors, with PVS volume increasing progressively across adulthood (Huang et al., 2021). In our study, we controlled for age, sex, and scanner effects, but we did not include measures of BMI, time of scanning, or cardiovascular parameters. These unmeasured variables could partly influence group differences between VP/VLBW and FT adults and should be considered in future research.

## Conclusion

In conclusion, our study demonstrates higher PVS volumes in basal ganglia and insula-related WM of VP/VLBW-born adults, with volumes linked to GA and INTI. These findings reveal a novel type of long-term structural alteration after VP/VLBW birth and contribute to a more complete understanding of the broad impact of prematurity on all relevant compartments of the brain. Given the importance of PVS for brain clearance, these alterations might have implications for long-term brain health in this vulnerable population.

## CRediT Author Statement

Niklas M. Hürter: Conceptualization, Methodology, Software, Validation, Formal analysis, Investigation, Data curation, Writing - original draft, Writing - review & editing, Visualization, Supervision

Vijay S. Schmenger: Conceptualization, Methodology, Software, Validation, Resources, Data curation

Taylor Barda: Validation, Investigation

Melissa Thalhammer: Writing - review & editing

Benita Schmitz-Koep: Data curation, Writing - review & editing, Supervision

Aurore Menegaux: Data curation, Writing - review & editing

Marcel Daamen: Writing - review & editing, Project administration

Henning Boecker: Writing - review & editing, Project administration

Josef Priller: Writing - review & editing

Andreas Decker: Writing - review & editing

Katerina Deike: Writing - review & editing

Claus Zimmer: Resources, Writing - review & editing

Peter Bartmann: Writing - review & editing, Project administration, Funding acquisition

Dieter Wolke: Writing - review & editing, Project administration, Funding acquisition Benedikt Zott: Writing - review & editing

Christian Sorg: Conceptualization, Resources, Writing - review & editing, Supervision, Project administration, Funding acquisition

Dennis M. Hedderich: Conceptualization, Writing - review & editing, Supervision, Project administration, Funding acquisition

## Data availability

The authors do not have permission to share data due to privacy restrictions and ethical considerations regarding participant confidentiality.

## Acknowledgments

This study was supported by the RECAP preterm (Research on European Children and Adults born Preterm) project, an EU Horizon 2020 study (Grant No. 733280 to D.W. and P.B.), the UK Research and Innovation (UKRI) Research Frontier Grant under the UK governments Horizon Europe funding guarantee (ERC-AdG) (grant number EP/X023206/1 to D.W.), the Deutsche Forschungsgemeinschaft (BA 6370/2-1 to C.S., HE 8967/3-1 to D.M.H., and ME 5894/2-1 to A.M.), the German Federal Ministry of Education and Science (BMBF 01ER0801 to P.B. and D.W., BMBF 01ER0803 to C.S.) and the Kommission für Klinische Forschung, Technische Universität München (KKF 8700000474 to D.M.H. and KKF 8700000811 to B.S.-K.).

## Declaration of generative AI and AI-assisted technologies in the writing process

During the preparation of this work Claude (Anthropic) was used to improve the linguistic quality and readability of the manuscript. This was limited to generating ideas and specific phrasings and did not involve creating entire text passages.

## Abbreviations

BLS: Bavarian Longitudinal Study
BW: Birth weight
CSF: Cerebrospinal fluid
DH: Duration of hospitalization
eTIV: Estimated total intracranial volume
FDR: False discovery rate
FT: Full-term
GA: Gestational age
INTI: Intensity of neonatal treatment index
IQ: Intelligence quotient
MANCOVA: Multivariate analysis of covariance
MRI: Magnetic resonance imaging
PVS: Perivascular spaces
VP/VLBW: Very preterm and/or very low birth weight
WM: White matter

