## Supplementary Material for "Higher perivascular space volume in very premature born adults"

Table of content

**List of Supplementary Figures**

**List of Supplementary Tables**

### **Supplementary Methods**

#### **Brain Parcellation**

Regional PVS analysis requires a consistent and anatomically meaningful subdivision of brain white matter. We used FreeSurfer's white matter parcellation (wmparc.mgz, version 7.3.2), which assigns white matter voxels to regional labels of cortical region parcellation based on the Desikan-Killiany atlas. Since wmparc provides region-specific rather than lobe-specific labels, individual wmparc codes were grouped into larger anatomical units (frontal, parietal, temporal, occipital, cingulate, insular, and unsegmented white matter, as well as basal ganglia) following standard neuroanatomical boundaries. The complete mapping of wmparc codes to these anatomical regions is provided in Table S1.

| Region | Included wmparc codes |
| --- | --- |
| Frontal Lobe | 3028, 4028: wm-lh/rh-superiorfrontal  3027, 4027: wm-lh/rh-rostralmiddlefrontal  3003, 4003: wm-lh/rh-caudalmiddlefrontal  3018, 4018: wm-lh/rh-parsopercularis  3020, 4020: wm-lh/rh-parstriangularis  3019, 4019: wm-lh/rh-parsorbitalis  3012, 4012: wm-lh/rh-lateralorbitofrontal  3014, 4014: wm-lh/rh-medialorbitofrontal  3024, 4024: wm-lh/rh-precentral  3017, 4017: wm-lh/rh-paracentral  3032, 4032: wm-lh/rh-frontalpole |
| Parietal Lobe | 3029, 4029: wm-lh/rh-superiorparietal  3008, 4008: wm-lh/rh-inferiorparietal  3031, 4031: wm-lh/rh-supramarginal  3022, 4022: wm-lh/rh-postcentral  3025, 4025: wm-lh/rh-precuneus |
| Temporal Lobe | 3030, 4030: wm-lh/rh-superiortemporal  3015, 4015: wm-lh/rh-middletemporal  3009, 4009: wm-lh/rh-inferiortemporal  3001, 4001: wm-lh/rh-bankssts  3007, 4007: wm-lh/rh-fusiform  3034, 4034: wm-lh/rh-transversetemporal  3006, 4006: wm-lh/rh-entorhinal  3033, 4033: wm-lh/rh-temporalpole  3016, 4016: wm-lh/rh-parahippocampal |
| Occipital Lobe | 3011, 4011: wm-lh/rh-lateraloccipital  3013, 4013: wm-lh/rh-lingual  3005, 4005: wm-lh/rh-cuneus  3021, 4021: wm-lh/rh-pericalcarine |
| Cingulate Cortex | 3026, 4026: wm-lh/rh-rostralanteriorcingulate  3002, 4002: wm-lh/rh-caudalanteriorcingulate  3023, 4023: wm-lh/rh-posteriorcingulate  3010, 4010: wm-lh/rh-isthmuscingulate |
| Insula | 3035, 4035: wm-lh/rh-insula |
| unsegmented | 5001, 5002: wm-lh/rh-unsegmented |
| Basal Ganglia | 10: Left-Thalamus-Proper  11: Left-Caudate  12: Left-Putamen  13: Left-Pallidum  49: Right-Thalamus-Proper  50: Right-Caudate  51: Right-Putamen  52: Right-Pallidum |
| Whole Brain | Regions 1-8 (all lobar white matter parcellations and Basal Ganglia) |
| White Matter | Regions 1-7 (lobar white matter parcellations only) |

Table S1: Individual white matter parcellation labels. Individual white matter parcellation labels from FreeSurfer's wmparc (Desikan-Killiany atlas, version 7.3.2) grouped into anatomical regions

#### **Definition of an Optimized PVS Segmentation Protocol**

The decisions to perform PVS analysis with for example nnU-Net-based segmentation or T2-weighted input MRIs in MNI space and at 1 mm³ isotropic resolution were not arbitrary but resulted from a systematic methodological comparison conducted prior to the main study. Below, we present the rationale and results of this methodological pre-study, which focused on two related questions: 1. which segmentation algorithm performs best to identify PVS in our data, and 2. which imaging input in terms of modality, resolution, and registration space yields the most reliable PVS segmentation.

##### **Comparison of Segmentation Algorithms**

We systematically evaluated five segmentation approaches on our dataset. Three were excluded as either the necessary training data were inaccessible or segmentation results proved inadequate: the Frangi filter-based approach (Frangi et al., 1998), the M2EDN approach (Lian et al., 2018), and the VALDO challenge methods approach (Sudre et al., 2024). WPSS (Lan et al., 2023) and nnU-Net (Isensee et al., 2021) alone yielded sufficient segmentation. Based on these results, we conducted a strict formal comparison between these two trainable deep learning approaches: the Weakly Supervised Perivascular Spaces Segmentation method (WPSS) and nnU-Net. WPSS integrates Frangi filter-derived geometric priors with CNN-based feature learning and was specifically designed for and validated exclusively on EPC images (Lan et al., 2023). Enhanced Perivascular Contrast (EPC) images, which are generated by voxel-wise division of T1-weighted by T2-weighted images, enhance PVS-to-tissue contrast and have been used to improve PVS segmentation (Sepehrband et al., 2019). Critically, the Frangi filter CNN within WPSS applies EPC-specific vesselness conditions (λ₂ ≤ 0, λ₃ ≤ 0) that rely on the inverted contrast profile of PVS in EPC images; the architecture cannot be applied to T2-weighted images without structural modification. WPSS was trained and validated on Human Connectome Project (HCP) data preprocessed using the HCP Minimal Processing Pipeline, which includes MNI152 registration (Sepehrband et al., 2019), meaning WPSS was itself developed and evaluated on MNI-normalized data. nnU-Net is a self-configuring general-purpose framework that has demonstrated state-of-the-art performance across 33 of 53 major medical image segmentation challenges without requiring domain-specific modifications.

Both networks were trained on 32 subjects and validated on 8 subjects, with both trained on EPC images to ensure comparability, consistent with the original WPSS design. WPSS produced substantial false-positive detections in sulcal and gray matter boundary regions; even after applying anatomical masks to remove these, WPSS consistently underperformed nnU-Net across all brain regions and metrics (Table S2).

| Region | Metric | nnU-Net | WPSS |
| --- | --- | --- | --- |
| Full brain | DSC | 0.527 | 0.381 |
|  | SEN | 0.451 | 0.342 |
|  | PPV | 0.652 | 0.445 |
| White matter | DSC | 0.493 | 0.370 |
|  | SEN | 0.412 | 0.340 |
|  | PPV | 0.636 | 0.428 |
| Basal ganglia | DSC | 0.623 | 0.367 |
|  | SEN | 0.604 | 0.362 |
|  | PPV | 0.665 | 0.436 |

Table S2: Segmentation performance metrics for WPSS and nnU-Net. Segmentation performance metrics for WPSS and nnU-Net relative to Rater 1's manual annotations, evaluated on a held-out validation set (n = 8 subjects). Both networks were trained on EPC images. DSC: Dice Similarity Coefficient; SEN: Sensitivity; PPV: Positive Predictive Value. nnU-Net outperforms WPSS across all regions and metrics.

##### **Comparison of MRI Input Data: Modality, Resolution, and Registration Space**

Having established nnU-Net as the superior algorithm, we evaluated the optimal imaging input with respect to modality and spatial configuration.

- Modality comparison: EPC vs. T2-weighted imaging. Enhanced Perivascular Contrast (EPC) images are generated by voxel-wise division of T1-weighted by T2-weighted images, thereby enhancing PVS-to-tissue contrast (Sepehrband et al., 2019). Since our T1-weighted images were acquired at 1 mm³ and T2-weighted images at 0.5 mm³ isotropic resolution, EPC generation required standardization of both modalities to 1 mm³. To ensure a methodologically fair modality comparison, T2-weighted images were registered to MNI space at the same 1 mm³ resolution. Two separate nnU-Net models were trained via five-fold cross-validation on all 40 subjects: one on EPC images (EPC-Net) and one on T2-weighted images (T2-Net) (Figure S1).

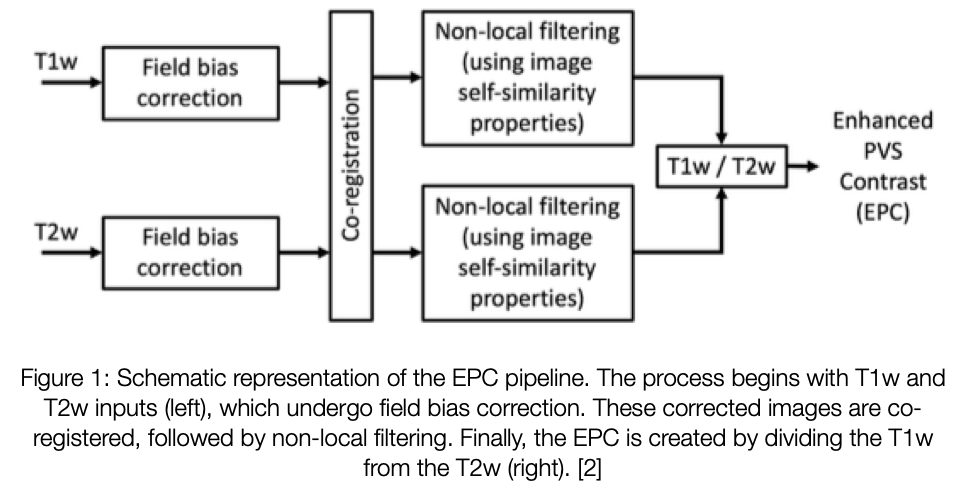

Figure S1: Schematic of the EPC pipeline. Enhanced Perivascular Contrast (EPC) images, which are generated by voxel-wise division of T1-weighted by T2-weighted images, enhance PVS-to-tissue contrast and have been used to improve PVS segmentation (Sepehrband et al., 2019).

T2-Net consistently outperformed EPC-Net across all brain regions and performance metrics (Table S3). The advantage was most pronounced in white matter and persisted in the basal ganglia. T2-Net also showed substantially smaller deviation from ground truth cluster counts (13.5% vs. 35.9% for EPC-Net). T2-weighted imaging was therefore selected as the input modality.

| Region | Metric | T2-Net | EPC-Net |
| --- | --- | --- | --- |
| Full brain | DSC | 0.637 | 0.504 |
|  | SEN | 0.556 | 0.421 |
|  | PPV | 0.771 | 0.678 |
| White matter | DSC | 0.611 | 0.477 |
|  | SEN | 0.525 | 0.393 |
|  | PPV | 0.764 | 0.665 |
| Basal ganglia | DSC | 0.716 | 0.595 |
|  | SEN | 0.665 | 0.539 |
|  | PPV | 0.791 | 0.702 |

Table S3: nnUnet segmentation performance metrics for T2 and EPC. Segmentation performance metrics for nnU-Net trained on T2-weighted (T2-Net) vs. EPC images (EPC-Net), evaluated using five-fold cross-validation (n = 40 subjects). DSC: Dice Similarity Coefficient; SEN: Sensitivity; PPV: Positive Predictive Value.

- Registration space and resolution: The use of MNI space for quantitative PVS analysis is an established practice in the published literature. Sepehrband et al. (Sepehrband et al., 2019) performed EPC-based PVS segmentation and quantification on MNI152-registered HCP data, and this pipeline has since been applied in several large-scale studies, including Sepehrband et al. (Sepehrband et al., 2021) and Lynch et al. (Lynch et al., 2023), the latter covering approximately 1,400 subjects across the lifespan. Ballerini et al. (Ballerini et al., 2018) additionally resampled MRI volumes to 1 mm isotropic voxels prior to applying Frangi filter-based PVS segmentation.

Our choice of MNI space over native space was determined by empirical comparison. When the nnU-Net model that was trained on MNI-normalized data with standardized spatial orientation and anatomical configuration was applied to native space data, qualitative review by Rater 1 confirmed markedly reduced segmentation sensitivity. Native space analysis additionally yielded reduced inter-rater specificity, despite theoretically preserving the original 0.5 mm³ resolution. This finding is consistent with the observations of Boutinaud et al. (Boutinaud et al., 2021), who tested MNI registration for a natively-trained PVS segmentation model and reported reductions in TPR (–15%) and PPV (–25%), attributing this to spatial standardization being incompatible with a model trained on native space data. MNI space standardization further ensures consistent anatomical correspondence across subjects, a prerequisite for the atlas-based regional white matter parcellation employed in this study. Importantly, registration-related effects apply equally to the VP/VLBW and full-term control groups, preserving the validity of between-group comparisons.

The 1 mm³ isotropic resolution reflects the EPC generation constraint described above (T1-weighted acquisition at 1 mm³) and was maintained for T2-weighted analysis to eliminate confounding effects of differing spatial configurations in the methodological comparison.

##### **Segmentation Performance in the Context of Published Literature**

The final T2-Net model achieved a mean DSC of 0.637 (range: 0.598–0.672), sensitivity of 0.556, and PPV of 0.771 across the full brain, based on five-fold cross-validation on all 40 training subjects. This exceeds the inter-rater DSC of 0.538 between Rater 1 and Rater 2 (Figure S2), demonstrating that the algorithm performs within the range of human rater variability. Regional performance was strongest in the basal ganglia (DSC = 0.716, SEN = 0.665, PPV = 0.791) compared to white matter (DSC = 0.611, SEN = 0.525, PPV = 0.764), a pattern consistent with the anatomical characteristics of PVS in these regions. Basal ganglia PVS are generally larger and less spatially dispersed, rendering them more accessible to automated segmentation.

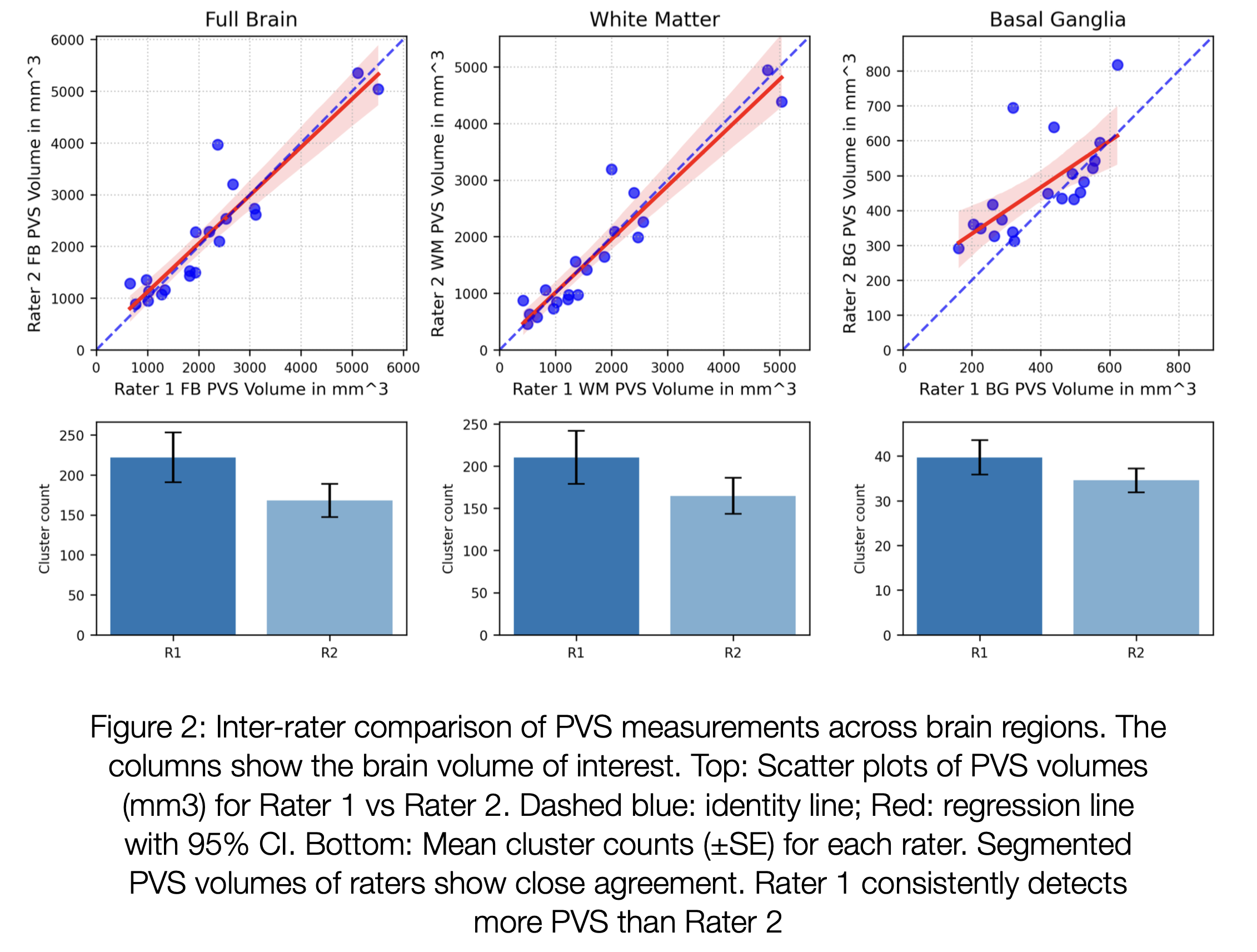

Figure S2: Inter-rater reliability of manual PVS segmentation. Columns show results for Full Brain (FB), White Matter (WM), and Basal Ganglia (BG). Top row: Scatter plots of absolute PVS volumes (mm³) annotated by Rater 1 vs. Rater 2 (n = 20 subjects); dashed blue line: identity line; solid red line: regression line with 95% CI. Bottom row: Mean PVS cluster counts (±SE) per rater. Volumes show close agreement between raters (ICC = 0.927); Rater 1 consistently detects a higher cluster count than Rater 2.

These results are in line with published benchmarks for automated PVS segmentation on 3T MRI (see Tables S4-8 below). In the VALDO challenge (Sudre et al., 2024) nnU-Net achieved DSC = 61.9%; our T2-Net achieves DSC = 63.7%. Cai et al. (Cai et al., 2024) reported DSC = 68% for nnU-Net on T2-weighted 3T data; our result is comparable, with the remaining difference attributable to dataset- and annotation-specific factors.

| **Descriptive Statistics of PVS Volume in our Study** | | | | |
| --- | --- | --- | --- | --- |
|  | Minimum | Maximum | Mean | Std. Deviation |
| Whole brain | .00040 | .00611 | .00217 | .00114 |
| WM | .00012 | .00577 | .00184 | .00119 |
| BG | .00173 | .01213 | .00661 | .00191 |

Table S4: Volume fraction (PVS volume divided by regional volume)

|  | Minimum | Maximum | Mean | Std. Deviation |
| --- | --- | --- | --- | --- |
| Whole brain | .0001442 | .0019279 | .0006917 | .0003597 |
| WM | .0000341 | .0016845 | .0005457 | .0003517 |
| BG | .0000335 | .0002632 | .0001460 | .0000433 |

Table S5: Volume fraction (PVS volume divided by eTIV volume)

|  | Minimum | Maximum | Mean | Std. Deviation |
| --- | --- | --- | --- | --- |
| Whole brain | 237 | 3114 | 1119.53 | 593.742 |
| WM | 54 | 2721 | 886.49 | 581.934 |
| BG | 64 | 406 | 233.05 | 62.231 |

Table S6: Absolute PVS volume (mm3)

We have systematically compared our results with the suggested reference studies (Barisano et al., 2021, Kim et al., 2023a) to address this observation.

| Study | Absolute Volume (mm³) | Fraction - ICV normalized (%) | Fraction - Regional normalized (%) |
| --- | --- | --- | --- |
| Our study | 886±582 | 0.055±0.035 | 0.184±0.119 |
| Kim et al. (2023) | 758±485 | 0.056±0.035 | Not reported |
| Barisano et al. (2021) | 5029±2,153 | Not reported | 1.14±0.43 |

Table S7: White Matter PVS Volume Comparison

| Study | Absolute Volume (mm³) | Fraction - ICV normalized (%) | Fraction - Regional normalized (%) |
| --- | --- | --- | --- |
| Our study | 233±62 | 0.015±0.004 | 0.661±0.191 |
| Kim et al. (2023) | 85±31 | 0.006±0.002 | Not reported |
| Barisano et al. (2021) | Not reported | Not reported | Not reported |

Table S8: Basal Ganglia PVS Volume Comparison

Our white matter PVS values closely match Kim et al. (886 vs. 758 mm³; 0.055% vs. 0.056% ICV-normalized), validating our technical approach. The substantially higher values reported by Barisano et al. (5029 mm³; 1.14% regional normalized fraction) are most likely explained by two methodological differences: their higher analytical resolution (native 0.7 mm³ vs. our 1 mm³ standardization), which preserves small PVS that may be lost through partial volume averaging in our pipeline, and their use of Frangi filtering, which is optimized for high sensitivity to tubular structures, compared to our conservative deep learning approach trained on manual annotations. Our elevated basal ganglia values relative to Kim et al.'s healthy controls (233 vs. 85 mm³) are biologically expected given our VP/VLBW cohort and are further supported by the significant correlations with gestational age and INTI. Critically, absolute volumetric calibration does not affect the internal validity of our between-group comparisons, which were conducted with consistent methodology applied equally to both groups.

To promote reproducibility, the trained model weights have been made publicly available (https://github.com/vijaysebastianS/nnU-Net-for-PVS-Segmentation-on-3T-T2w-MRI).

### **Supplementary Results**

#### **Control for normalization approaches in PVS volume group comparisons**

All three MANCOVA analyses yielded significant multivariate group effects (same region normalized: Wilks' λ = .789, p < .001, partial η² = .211; eTIV normalized: Wilks' λ = .909, p = .031, partial η² = .091; absolute volumes with eTIV covariate: Wilks' λ = .914, p = .046, partial η² = .086).

| **Normalization approach** | **Region** | **VP/VLBW Mean (SD)** | **Full-Term Mean (SD)** | **Partial Eta Squared** | **p-value** | **FDR cut-off** |
| --- | --- | --- | --- | --- | --- | --- |
| *Same region volume normalized* | whole brain | 0.00230 (0.00122) | 0.00199 (0.00095) | 0.020 | 0.056 | 0.02 |
|  | whole WM | 0.00195 (0.00128) | 0.00168 (0.00100) | 0.013 | 0.121 | 0.03 |
|  | frontal WM | 0.00230 (0.00185) | 0.00191 (0.00155) | 0.012 | 0.132 | 0.035 |
|  | parietal WM | 0.00279 (0.00197) | 0.00249 (0.00146) | 0.007 | 0.250 | 0.045 |
|  | temporal WM | 0.00042 (0.00052) | 0.00038 (0.00038) | 0.002 | 0.599 | 0.05 |
|  | occipital WM | 0.00010 (0.00016) | 0.00007 (0.00011) | 0.010 | 0.180 | 0.04 |
|  | cingulate WM | 0.00065 (0.00086) | 0.00042 (0.00057) | 0.023 | 0.039* | 0.015 |
|  | insula WM | 0.00294 (0.00183) | 0.00216 (0.00132) | 0.057 | 0.001** | 0.01 |
|  | unsegmented WM | 0.00323 (0.00196) | 0.00279 (0.00159) | 0.015 | 0.094 | 0.025 |
|  | basal ganglia | 0.00713 (0.00200) | 0.00600 (0.00157) | 0.096 | <0.001** | 0.005 |
| *eTIV normalized* | whole brain | 0.000720 (0.000379) | 0.000640 (0.000310) | 0.013 | 0.120 | 0.02 |
|  | whole WM | 0.000570 (0.000375) | 0.000500 (0.000302) | 0.009 | 0.191 | 0.03 |
|  | frontal WM | 0.000220 (0.000176) | 0.000180 (0.000153) | 0.011 | 0.162 | 0.035 |
|  | parietal WM | 0.000170 (0.000122) | 0.000150 (0.000086) | 0.008 | 0.228 | 0.045 |
|  | temporal WM | 0.0000173 (0.0000214) | 0.0000159 (0.0000158) | 0.001 | 0.612 | 0.05 |
|  | occipital WM | 0.0000029 (0.0000048) | 0.0000020 (0.0000034) | 0.012 | 0.149 | 0.04 |
|  | cingulate WM | 0.0000104 (0.0000144) | 0.0000068 (0.0000092) | 0.022 | 0.047* | 0.015 |
|  | insula WM | 0.0000354 (0.0000216) | 0.0000264 (0.0000167) | 0.052 | 0.002** | 0.01 |
|  | unsegmented WM | 0.000120 (0.0000757) | 0.000120 (0.0000714) | 0.000 | 0.873 | 0.025 |
|  | basal ganglia | 0.000150 (0.0000455) | 0.000137 (0.0000383) | 0.030 | 0.018* | 0.005 |
| *Absolute volumes with eTIV as covariate* | whole brain | 1172.703 (616.169) | 1039.177 (531.688) | 0.012 | 0.135 | 0.02 |
|  | whole WM | 936.458 (610.269) | 811.411 (518.871) | 0.011 | 0.154 | 0.03 |
|  | frontal WM | 351.254 (274.116) | 299.656 (256.642) | 0.008 | 0.216 | 0.035 |
|  | parietal WM | 273.436 (195.931) | 249.334 (144.021) | 0.004 | 0.373 | 0.045 |
|  | temporal WM | 28.461 (35.005) | 25.172 (25.468) | 0.003 | 0.496 | 0.05 |
|  | occipital WM | 5.000 (8.085) | 3.247 (5.661) | 0.014 | 0.107 | 0.04 |
|  | cingulate WM | 18.204 (24.568) | 10.238 (16.266) | 0.033 | 0.014** | 0.015 |
|  | insula WM | 58.221 (34.243) | 40.950 (26.531) | 0.069 | <0.001** | 0.01 |
|  | unsegmented WM | 201.882 (135.695) | 182.814 (131.161) | 0.006 | 0.302 | 0.025 |
|  | basal ganglia | 236.245 (64.979) | 227.766 (56.393) | 0.005 | 0.369 | 0.005 |

Table S9: PVS volume differences in different normalization approaches. * indicates p<0.05; ** indicates p_FDR_<0.05.

#### **Hemispheric differences in significant regions**

The general linear model analysis examined hemispheric differences in PVS volumes between VP/VLBW and control groups across three brain regions (cingulate, insula, and basal ganglia), while controlling for sex, age at scan, and scanner. Significant bilateral differences were observed in the insula (right: p=0.026; left: p=0.001) and basal ganglia (right: p=<0.001; left: p=0.001), with VP/VLBW-born participants consistently showing larger PVS volumes than controls. In the cingulate region, only the right hemisphere showed a significant difference (p=0.019), while the left hemisphere difference did not reach statistical significance (p=0.166). The largest effect sizes were found in the right basal ganglia (partial η²=0.082), left insula (partial η²=0.060), and left basal ganglia (partial η²=0.056), all with high statistical power (>0.900).

| *Region* | *VP/VLBW Mean (SD)* | *Full-Term Mean (SD)* | *p-value* | *Partial Eta Squared* |
| --- | --- | --- | --- | --- |
| *cingulate WM right* | 0.00071 (0.00104) | 0.00040 (0.00064) | 0.019 | 0.030 |
| *cingulate WM left* | 0.00059 (0.00085) | 0.00043 (0.00066) | 0.166 | 0.011 |
| *insula WM right* | 0.00282 (0.00201) | 0.00223 (0.00148) | 0.026 | 0.027 |
| *insula WM left* | 0.00307 (0.00217) | 0.00210 (0.00164) | <0.001 | 0.060 |
| *basal ganglia right* | 0.00726 (0.00263) | 0.00595 (0.00180) | <0.001 | 0.082 |
| *basal ganglia left* | 0.00701 (0.00219) | 0.00605 (0.00183) | 0.001 | 0.056 |

Table S10: Hemispheric regional PVS differences.

#### **Correlation with birth variables and IQ**

The Spearman’s correlation revealed a significant negative correlation between gestational age and basal ganglia PVS volume (ρ =-0.223, p=0.030) and a significant positive correlation between INTI and basal ganglia PVS volume (ρ =0.222, p=0.030). Birth weight and duration in hospital showed no significant correlations with PVS volumes.

| Variable 1 | Variable 2 | Spearman's Rho | p-value |
| --- | --- | --- | --- |
| whole brain | Gestational age | 0.013 | 0.899 |
|  | Birth weight | -0.124 | 0.234 |
|  | Duration in hospital | -0.006 | 0.957 |
|  | INTI | 0.052 | 0.613 |
| whole WM | Gestational age | 0.042 | 0.688 |
|  | Birth weight | -0.118 | 0.255 |
|  | Duration in hospital | -0.009 | 0.93 |
|  | INTI | 0.043 | 0.675 |
| frontal WM | Gestational age | 0.088 | 0.398 |
|  | Birth weight | -0.102 | 0.328 |
|  | Duration in hospital | -0.026 | 0.804 |
|  | INTI | 0.03 | 0.771 |
| parietal WM | Gestational age | -0.002 | 0.986 |
|  | Birth weight | -0.169 | 0.104 |
|  | Duration in hospital | 0.03 | 0.773 |
|  | INTI | 0.063 | 0.542 |
| temporal WM | Gestational age | 0.123 | 0.239 |
|  | Birth weight | -0.093 | 0.372 |
|  | Duration in hospital | -0.076 | 0.468 |
|  | INTI | -0.094 | 0.361 |
| occipital WM | Gestational age | 0.071 | 0.496 |
|  | Birth weight | 0.169 | 0.104 |
|  | Duration in hospital | -0.124 | 0.235 |
|  | INTI | -0.032 | 0.754 |
| cingulate WM | Gestational age | 0.045 | 0.666 |
|  | Birth weight | 0.001 | 0.992 |
|  | Duration in hospital | -0.098 | 0.35 |
|  | INTI | 0.065 | 0.529 |
| insula WM | Gestational age | 0.113 | 0.276 |
|  | Birth weight | -0.025 | 0.808 |
|  | Duration in hospital | -0.126 | 0.225 |
|  | INTI | 0.089 | 0.39 |
| unsegmented WM | Gestational age | -0.079 | 0.448 |
|  | Birth weight | -0.122 | 0.242 |
|  | Duration in hospital | 0.019 | 0.855 |
|  | INTI | 0.035 | 0.731 |
| basal ganglia | Gestational age | -0.223 | 0.030* |
|  | Birth weight | -0.18 | 0.082 |
|  | Duration in hospital | 0.138 | 0.184 |
|  | INTI | 0.222 | 0.030* |

Table S11: Regional PVS volume correlation with birth variables

Regarding cognitive outcomes, the Spearman's rho correlation analysis showed significant positive correlations between Full-Scale IQ and PVS volumes in some brain regions. Nevertheless, there was no significant correlation in basal ganglia or insula.

| Variable 1 | Variable 2 | Spearman's Rho | p-value |
| --- | --- | --- | --- |
| whole brain | Full-Scale IQ | 0.236 | 0.022 * |
| whole WM | Full-Scale IQ | 0.224 | 0.030* |
| frontal WM | Full-Scale IQ | 0.208 | 0.044* |
| parietal WM | Full-Scale IQ | 0.16 | 0.124 |
| temporal WM | Full-Scale IQ | 0.243 | 0.018* |
| occipital WM | Full-Scale IQ | 0.419 | <0.001* |
| cingulate WM | Full-Scale IQ | 0.198 | 0.056 |
| insula WM | Full-Scale IQ | 0.17 | 0.102 |
| unsegmented WM | Full-Scale IQ | 0.177 | 0.088 |
| basal ganglia | Full-Scale IQ | -0.047 | 0.653 |

Table S12: Regional PVS volume correlation with IQ

#### **PVS count analysis**

PVS count analysis revealed significant differences between VP/VLBW and FT participants in three specific brain regions: basal ganglia (mean difference=0.000108, p<0.001, partial η²=0.060), insula-related WM (mean difference=0.000138, p=0.010, partial η²=0.036), and cingulate-related WM (mean difference=0.000052, p=0.030, partial η²=0.026). Further hemispheric analysis demonstrated significant bilateral differences in the basal ganglia (right: p=0.001, partial η²=0.057; left: p=0.003, partial η²=0.047) and insula WM (right: p=0.019, partial η²=0.030; left: p=0.008, partial η²=0.038). In the cingulate region, only the right hemisphere showed a significant difference (p=0.014, partial η²=0.033), while the left hemisphere difference did not reach statistical significance (p=0.092). The remaining brain regions showed no significant differences in normalized PVS count between groups (all p>0.05).

| *Dependent Variable* | *VP/VLBW Mean (SD)* | *Full-Term Mean (SD)* | *F Value* | *p-value* | *Partial Eta Squared* | *Observed Power* |
| --- | --- | --- | --- | --- | --- | --- |
| *whole brain* | 0.0003182 (0.0001509) | 0.0003115 (0.0001368) | 0.205 | 0.651 | 0.001 | 0.074 |
| *whole WM* | 0.0003116 (0.0001605) | 0.0003069 (0.0001482) | 0.118 | 0.731 | 0.001 | 0.064 |
| *frontal WM* | 0.0004128 (0.0002550) | 0.0004118 (0.0002601) | 0.091 | 0.764 | 0.001 | 0.060 |
| *parietal WM* | 0.0005045 (0.0003003) | 0.0005144 (0.0002791) | 0.082 | 0.776 | 0.000 | 0.059 |
| *temporal WM* | 0.0001064 (0.0001137) | 0.0001078 (0.0001016) | 0.090 | 0.765 | 0.000 | 0.060 |
| *occipital WM* | 0.0000327 (0.0000469) | 0.0000290 (0.0000439) | 0.713 | 0.399 | 0.004 | 0.134 |
| *cingulate WM* | 0.0001620 (0.0001877) | 0.0001164 (0.0001212) | 4.781 | 0.030* | 0.026 | 0.585 |
| *insula WM* | 0.0007458 (0.0003904) | 0.0006188 (0.0003199) | 6.689 | 0.010* | 0.036 | 0.730 |
| *unsegmented WM* | 0.0007868 (0.0003858) | 0.0007185 (0.0003315) | 1.669 | 0.198 | 0.009 | 0.250 |
| *basal ganglia* | 0.0008305 (0.0002147) | 0.0007272 (0.0002151) | 11.615 | <0.001* | 0.060 | 0.924 |

Table S13: Region-normalized PVS count group differences between VP/VLBW and FT

| Dependent Variable | Mean Difference | p-value | F value | Partial Eta Squared | Observed Power |
| --- | --- | --- | --- | --- | --- |
| cingulate WM right | 0.0000612 | 0.014* | 6.127 | 0.033 | 0.693 |
| cingulate WM left | 0.0000432 | 0.092 | 2.872 | 0.016 | 0.392 |
| insula WM right | 0.0001254 | 0.019* | 5.612 | 0.030 | 0.655 |
| insula WM left | 0.0001506 | 0.008* | 7.227 | 0.038 | 0.763 |
| basal ganglia right | 0.0001212 | 0.001* | 10.994 | 0.057 | 0.911 |
| basal ganglia left | 0.0000948 | 0.003* | 9.047 | 0.047 | 0.849 |

significant = *

Table S14: Hemispheric region-normalized PVS count group differences.
